# Genetic associations for two biological age measures point to distinct aging phenotypes

**DOI:** 10.1101/2020.07.10.20150797

**Authors:** Chia-Ling Kuo, Luke C. Pilling, Zuyun Liu, Janice L. Atkins, Morgan E. Levine

**Affiliations:** Connecticut Convergence Institute for Translation in Regenerative Engineering, University of Connecticut Health, Farmington, Connecticut, USA.; University of Connecticut Center on Aging, School of Medicine, Farmington, Connecticut, USA.; College of Medicine and Health, University of Exeter, UK.; Department of Big Data in Health Science, School of Public Health and the Second Affiliated Hospital, Zhejiang University School of Medicine, Hangzhou, Zhejiang, China; Department of Pathology, Yale School of Medicine, New Haven, Connecticut, USA.; Department of Epidemiology, Yale School of Public Health, New Haven, Connecticut, USA

**Author notes:** Corresponding authors: Morgan Levine, 300 George St., New Haven, CT 06519, Chia-Ling Kuo, 195 Farmington Ave, Suite 2080, Farmington, CT 06032.

**Keywords:** *APOE*, biomarkers, polygenic risk score, cardiac aging, inflammaging

## Abstract

Biological age measures outperform chronological age in predicting various aging outcomes, yet little is known regarding genetic predisposition. We performed genome-wide association scans of two age-adjusted biological age measures (PhenoAgeAcceleration and BioAgeAcceleration), estimated from clinical biochemistry markers^1,2^ in European-descent participants from UK Biobank. The strongest signals were found in the *APOE* gene, tagged by the two major protein-coding SNPs, PhenoAgeAccel—rs429358 (*APOE* e4 determinant) (p=1.50×10^−72^); BioAgeAccel—rs7412 (*APOE* e2 determinant) (p=3.16×10^−60^). Interestingly, we observed inverse *APOE* e2 and e4 associations and unique pathway enrichments when comparing the two biological age measures. Genes associated with BioAgeAccel were enriched in lipid related pathways, while genes associated with PhenoAgeAccel showed enrichment for immune system, cell function, and carbohydrate homeostasis pathways, suggesting the two measures capture different aging domains. Our study reaffirms that aging patterns are heterogenous across individuals, and the manner in which a person ages may be partly attributed to genetic predisposition.

## Introduction

While chronological age is arguably the strongest determinant of risk for major chronic diseases, it alone is not sufficient to reflect the state of biological aging. Individuals with similar chronological ages are heterogenous in their physiological states, and subsequent health risks, due to differences in both the rate and manner of biological aging. As a result, efforts have been launched to develop measures that can capture the concept of biological age (BA)^3^. Typically, these measures encompass single or composite biomarkers found to be associated with a surrogate of biological age, usually chronological age or mortality. In principle, a valid BA measure needs to outperform chronological age in predicting lifespan and a wide range of age-sensitive tests in multiple physiological and behavioral domains^4^. BA predictors may help discover drivers of the aging process and can be used for secondary prevention by identifying at-risk individuals prior to disease onset. Additionally, they have been proposed as tools to monitor intervention or treatment effects aimed at targeting the aging process^5^.

A variety of data modalities, covering different biological levels of organization, have been used to develop BA predictors, e.g., DNA methylation data, gene expression data, proteomic data, metabolomic data, and clinical chemistry measures^6,7^. A study comparing 11 BA predictors found low agreement between those based on DNA methylation, clinical biomarkers, and telomere length, in terms of both their correlations with each other and their relative associations with healthspan-related characteristics: balance, grip strength, motor coordination, physical limitations, cognitive decline, self-rated health, and facial aging^8^. These findings suggest that different BA predictors may be capturing distinct aspects of the aging process. Furthermore, a recent paper^9^ combining various omics data identified distinct “ageotypes”, which represent diverse aging patterns across individuals.

We hypothesize that individual susceptibility to one biological aging domain versus another may be due in part to underlying genetic mechanisms. To test the hypothesis, we used data from the UK Biobank study to understand genetic predisposition to accelerated aging, measured by two validated biological age predictors (PhenoAge^1^ and BioAge^2^). PhenoAge is a function of chronological age, albumin, creatinine, C-reactive protein (CRP), alkaline phosphatase, glucose, lymphocyte percentage, mean corpuscular volume, red blood cell distribution width (RDW), and white blood cell count. BioAge is function of chronological age, albumin, creatinine, CRP, and alkaline phosphatase (also in PhenoAge), plus glycated hemoglobin (HbA1c), systolic blood pressure, and total cholesterol. Both aging measures have been shown to be robust predictors of aging outcomes^1,2,10^, yet are clearly distinct. Biological age acceleration measured by either biological age measure adjusted for chronological age (PhenoAgeAccel or BioAgeAccel) was similarly associated with morbidity and mortality in the full sample and subgroups of National Health and Nutrition Examination Survey (NHANES) IV, but PhenoAgeAccel outperformed BioAgeAccel in those disease-free and with normal body mass index^10^. Overall, this study is fully supported by UK Biobank, featured by a large sample and extensive genetic and phenotypic data.

## Results

451,367 genetically-determined Europeans were identified in UK Biobank. Of whom, 379,703 unrelated participants were included in analyses. Among the included samples (Table 1), 204,736 (54%) participants were female. After a mean follow-up time of 11.49 years (standard deviation (SD)=1.55) to April 26, 2020 (last death in the data), 23,060 participants died with the mean age at death 69.06 years (SD=7.21, range: 40.84 to 82.50). A summary of PhenoAge or BioAge biomarkers is provided in Table 1.

**Table 1.**
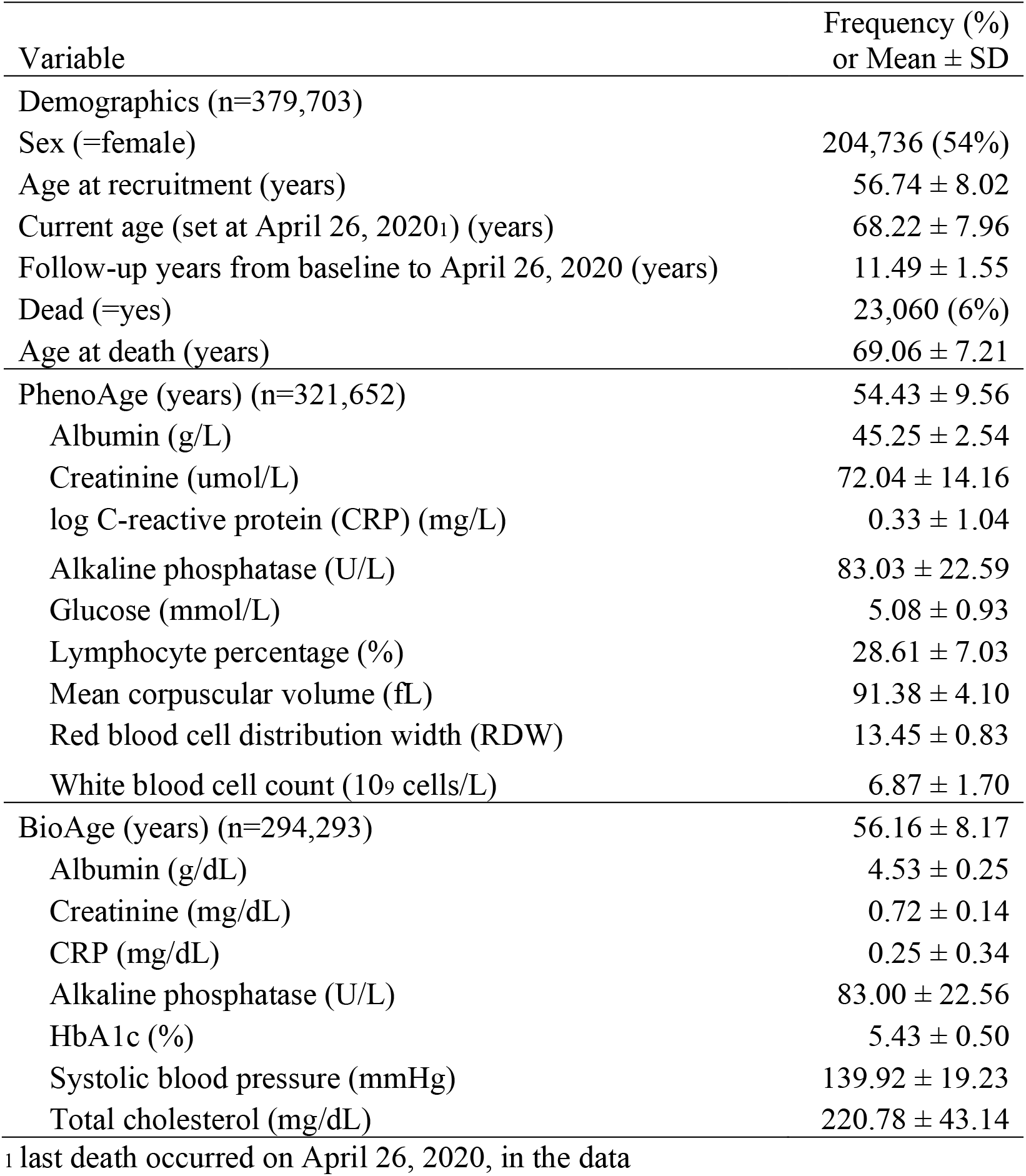
A summary for demographics and biomarkers in PhenoAge or BioAge

Participants were biologically younger than their chronological ages, with the mean PhenoAge and BioAge, 54.43 years (SD=9.56) and 56.16 years (SD=8.17) versus the mean chronological age 56.74 years (SD=8.02). PhenoAge acceleration (PhenoAgeAccel) estimated by residualizing PhenoAge based on chronological age via a linear regression model was weakly correlated (r=0.23) with BioAge acceleration (BioAgeAccel) that was similarly defined. Both BioAgeAccel and PhenoAgeAccel were significantly associated with all-cause mortality in this young cohort (p<2×10^−6^), with the hazard ratio (HR) 1.100 (95% CI: 1.097 to 1.102) per year increase in PhenoAgeAccel and 1.054 (95% CI: 1.046 to 1.062) per year increase in BioAgeAccel. In the above models, sex was included, additional to chronological age and PhenoAge or BioAge. When both PhenoAge and BioAge were included, the hazard ratio with PhenoAgeAccel was little changed (HR=1.099, 95% CI: 1.097 to 1.102) but that with BioAgeAccel (HR=1.000, 95% CI: 0.992 to 1.008) was reduced towards the null.

The data was split with a 1 to 2 ratio, where PhenoAge was available for 107,460 participants in the training set and for 214,192 participants in the testing set. Similarly, BioAge was available for 98,446 participants in the training set and for 195,847 participants in the testing set. The training set was used to perform genome-wide association analysis and the GWAS summary statistics were used to construct polygenic risk scores (PRSs) in the testing set, evaluated for the use of risk stratification for age-related outcomes. Demographics and PhenoAge or BioAge biomarker levels were quite balanced between the training and testing sets (Table S1).

### PhenoAgeAccel GWAS

In the Genome-Wide Association Study (GWAS) of PhenoAgeAccel, 7,561 genetic variants were identified (p<5×10^−8^) (Figure 1). The SNP p-value distribution showed sizable deviation from the null distribution of no association. However, there was a lack of evidence of population stratification or cryptic relatedness (LD score regression intercept 1.02 with the standard error (SE) 0.01, compared to the null value 1), and the proportion of inflation not explained by polygenic heritability was only 6.33% (SE=3.06%).

**Figure 1.**
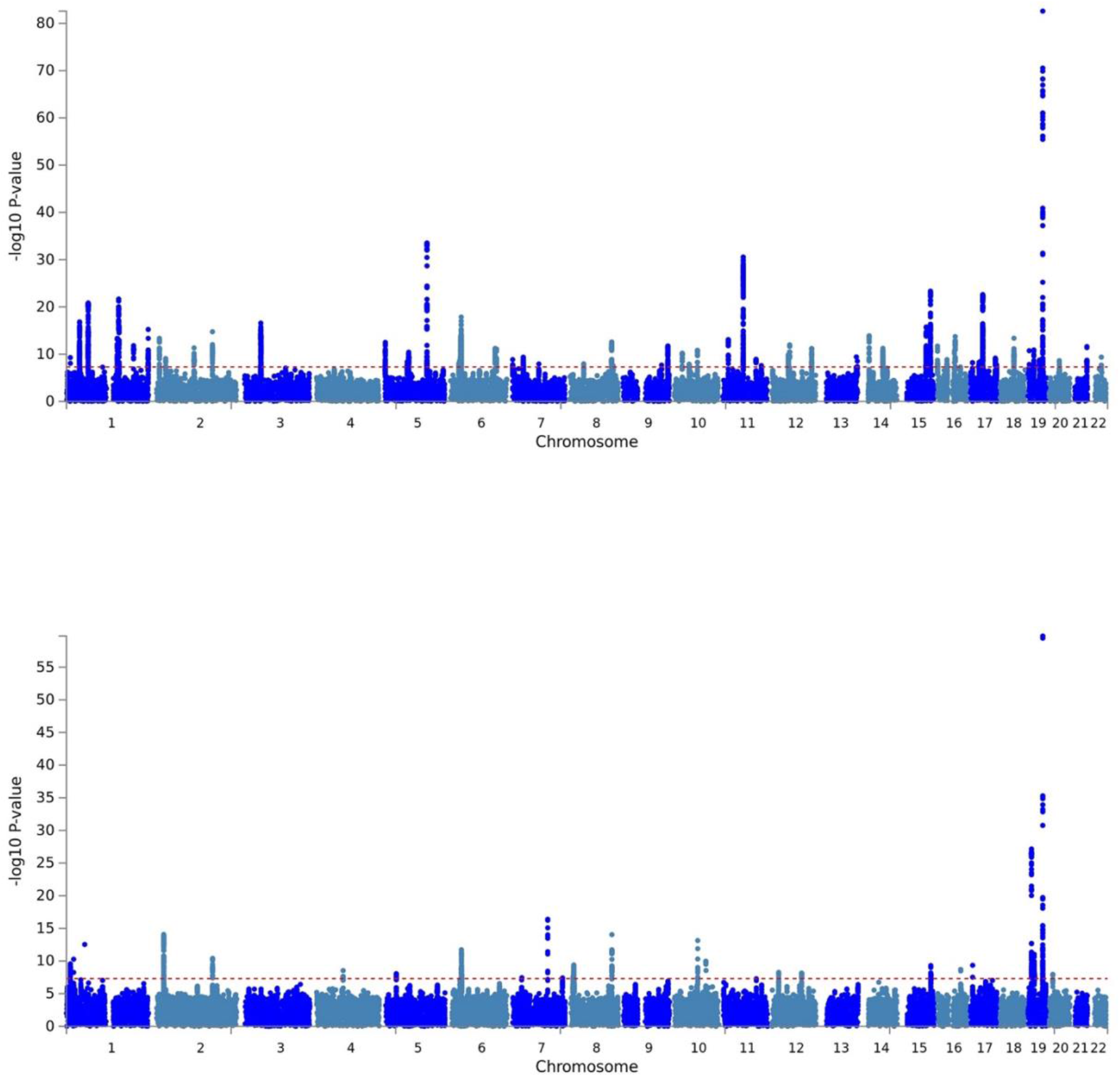
PhenoAgeAccel (top) and BioAgeAccel (bottom) Manhattan plots.

The SNP-heritability for PhenoAgeAccel was estimated to be 14.45% (SE=0.95%). We identified 55 independent signals (p<5×10^−8^) tagged by 55 lead SNPs. Of which, 29 were near genes (Table 2). Both *APOE* isoform coding SNPs (rs429358 and rs7412 on chromosome 19) were identified. Multiple lead SNPs were associated with CRP, glucose or HbA1c, and hematology traits, based on previous GWAS catalog^11^ results (Table S2). Lead SNPs nearby *GCKR, FTO, ZPR1*, and *APOE* were associated with various traits including cardiovascular diseases and/or PhenoAge biomarkers (Table S2).

**Table 2.**
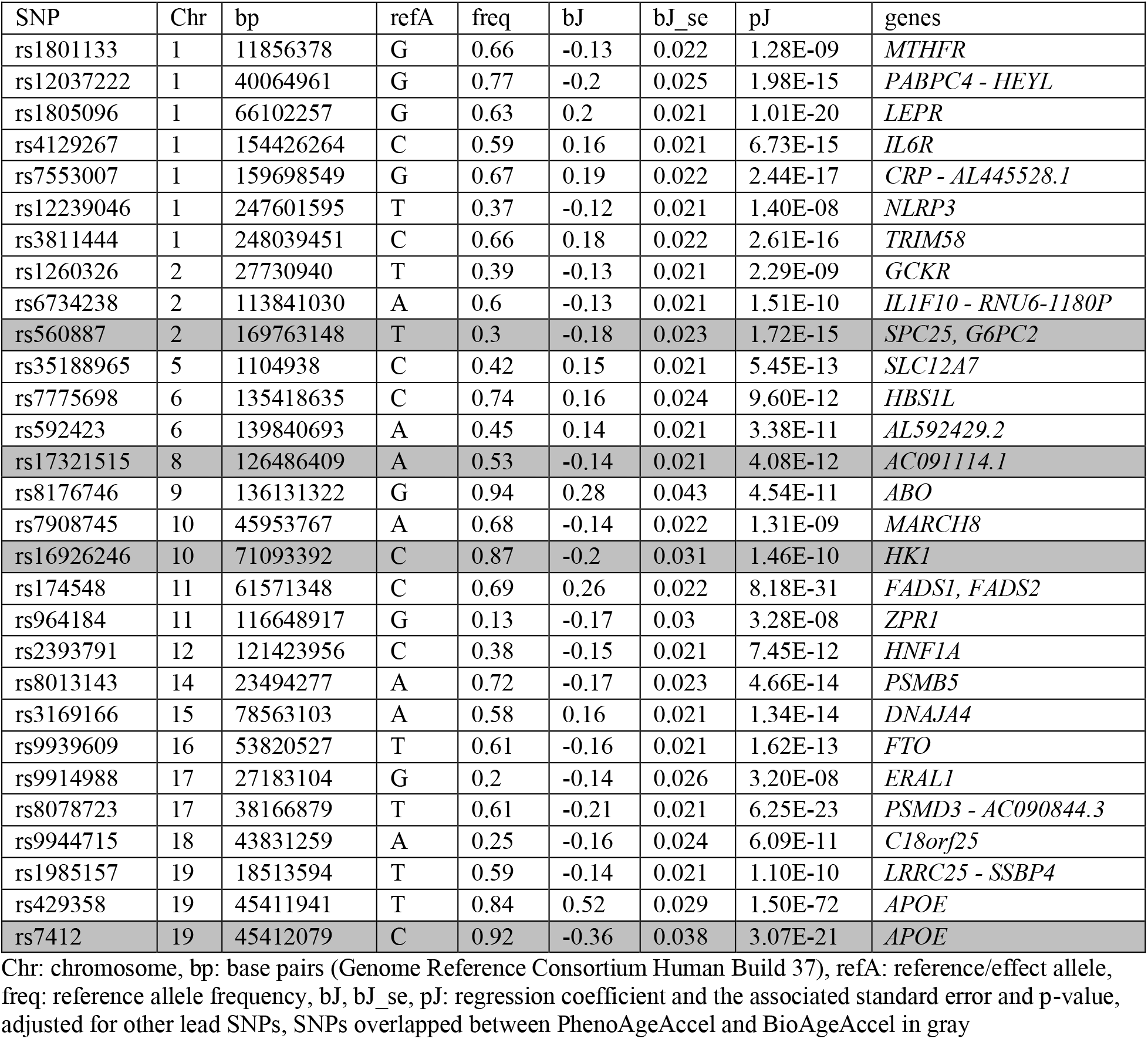
Genetic loci associated with PhenoAgeAccel (p<5⨯10^−8^) that can be mapped to genes

The Multi-marker Analysis of GenoMic Annotation (MAGMA) gene set analysis identified 11 gene sets at the Bonferroni-corrected level of 5%, including regulation of signaling and transcription, homeostasis (carbohydrate homeostasis, homeostasis of number of cells, and myeloid cell homeostasis), and immune system process (Figure 2). In the MAGMA tissue expression analysis, we found that genes expressed in whole blood and liver were more likely to be associated with PhenoAgeAccel than genes expressed in other tissues (Figure 3).

**Figure 2.**
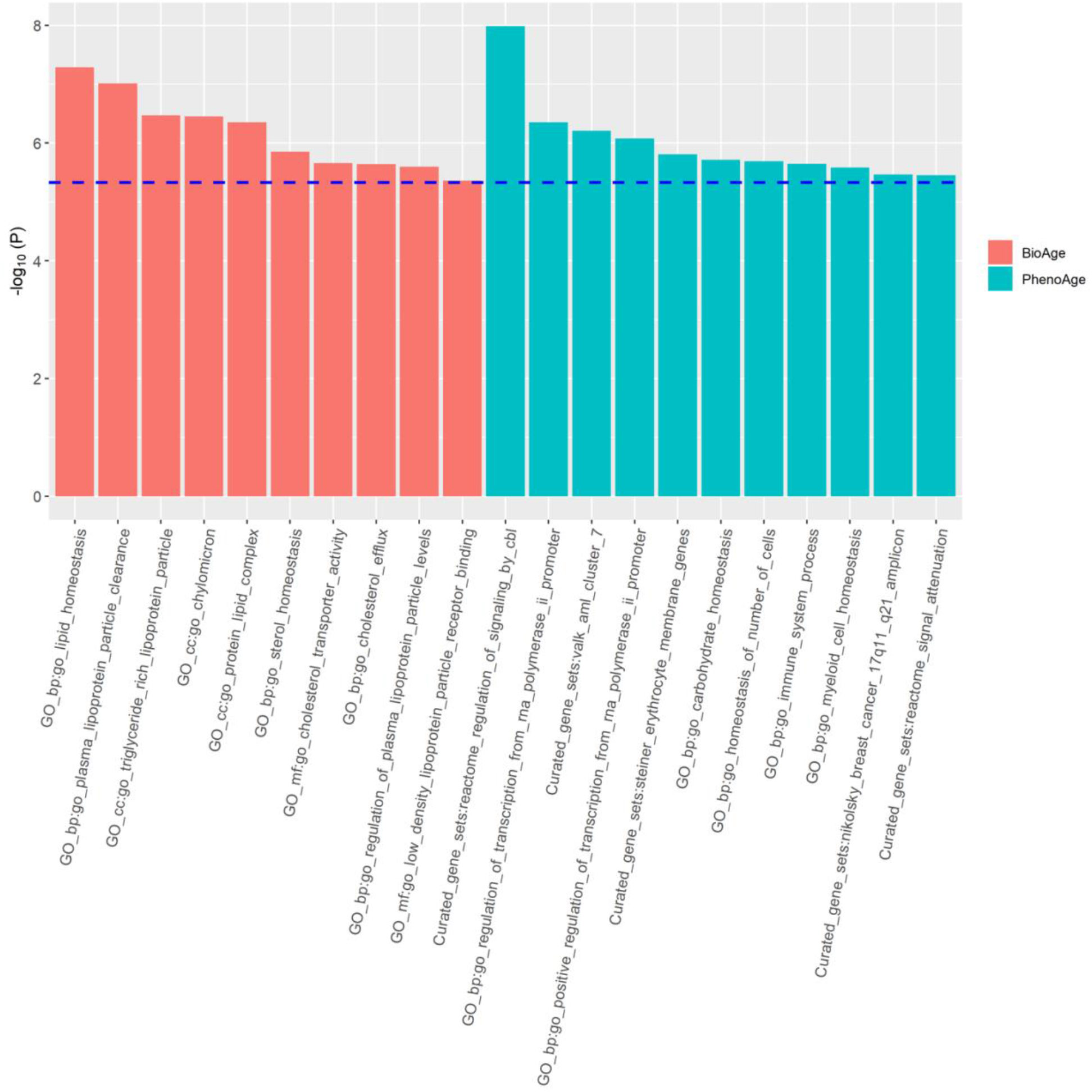
Significant gene sets identified by MAGMA for PhenoAgeAccel and BioAgeAccel.

**Figure 3.**
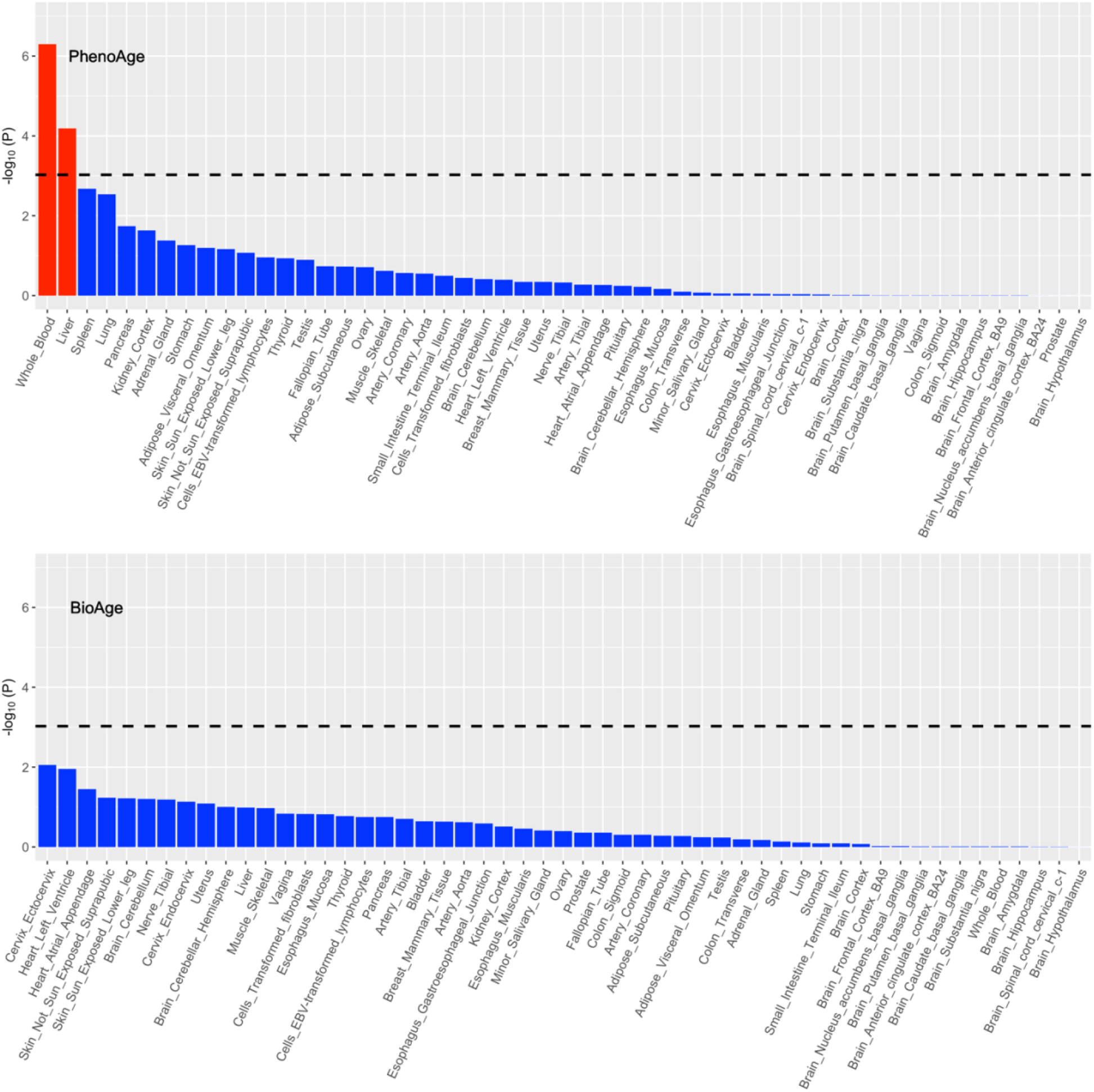
Association between tissue-specific gene expression and PhenoAgeAccel-gene or BioAgeAccel-gene association.

### BioAgeAccel GWAS

In the GWAS for BioAgeAccel, 996 genetic variants were identified (p<5×10^−8^) (Figure 1). The observed p-value distribution was significantly deviant from the expected under the null (Figure 2). However, there was no evidence to suggest population stratification or cryptic relatedness (LD score regression intercept=1.02, SE=0.01), and the proportion of inflation not explained by polygenic heritability was small, 6.58% (SE=3.78%).

The SNP-heritability for BioAgeAccel was estimated to be 12.39% (SE=0.95%). Twenty lead SNPs were identified (p<5×10^−8^) and 16 were nearby genes (Table 3). The strongest signal appeared in the *APOE* gene, tagged by the *APOE* isoform coding SNP rs7412. Several lead SNPs were associated with blood pressures. Other lead SNPs were associated with HbA1c, cardiovascular disease, and/or lipid biomarkers (Table S3).

**Table 3.**
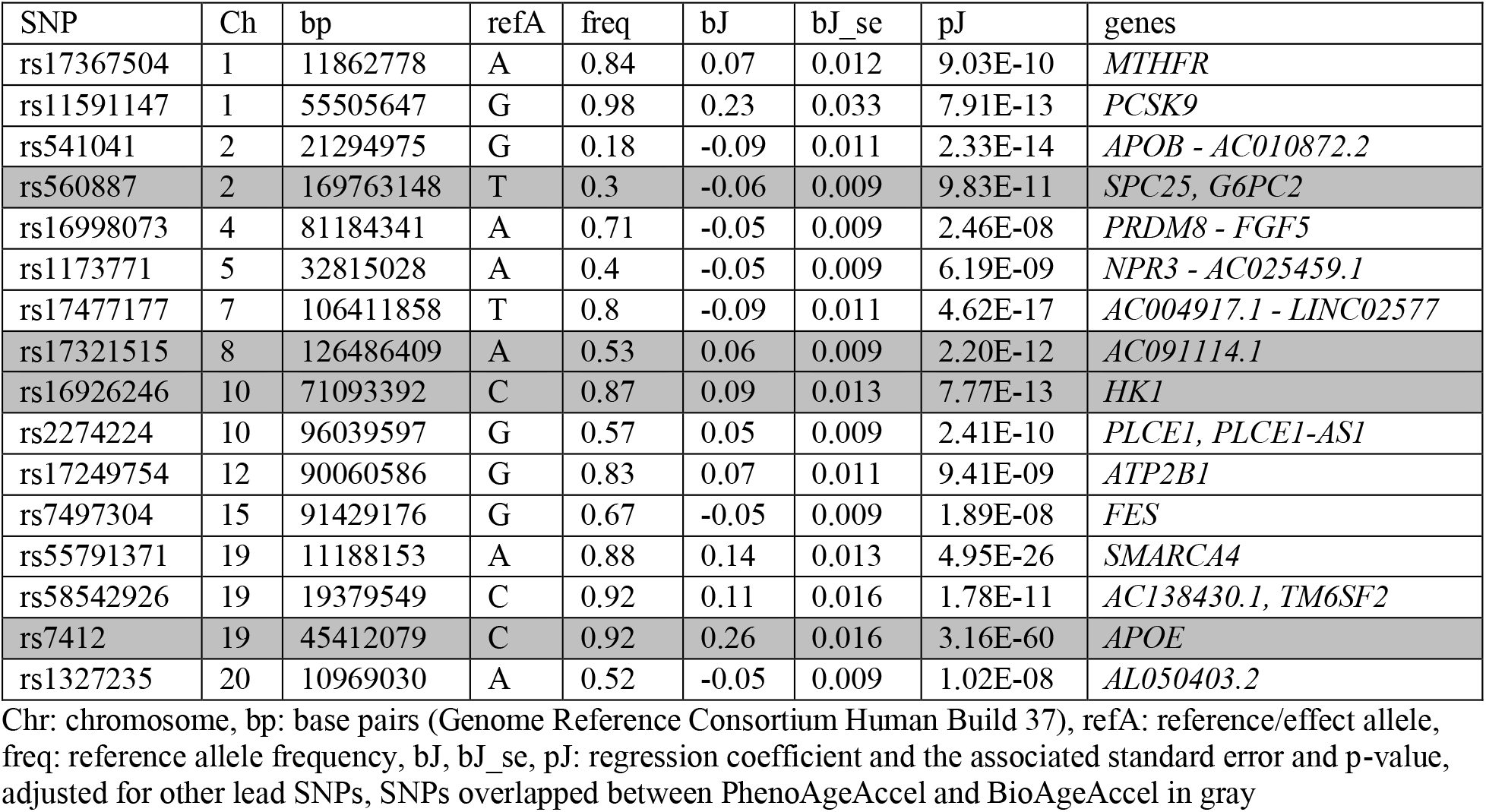
Genetic loci associated with BioAgeAccel (p<5⨯10^−8^) that can be mapped to genes

The MAGMA gene set analysis identified 10 lipid-related gene sets at the Bonferroni-corrected level of 5%, including lipid homeostasis, lipid protein particle clearance, and triglyceride-rich plasma lipoprotein particle (Figure 2). None of the 53 tissues showed significant specificity in gene expression for the genes associated with BioAgeAccel (Figure 3).

### PhenoAgeAccel GWAS vs. BioAgeAccel GWAS

PhenoAgeAccel and BioAgeAccel shared the lead SNPs, rs560887 (near *SPC25, G6PC2*), rs17321515 (near *AC091114.1*), rs16926246 (near *HK1*), and rs7412 (near *APOE*). Interestingly, three out of four common lead SNPs were oppositely associated with PhenoAgeAccel and BioAgeAccel.

- The rs560887 T allele, associated with decreased HbA1c^12,13^, is associated with decreased BioAgeAccel and PhenoAgeAccel.
- The rs16926246 C allele, associated with increased HbA1c^14^, is associated with increased BioAgeAccel, but decreased PhenoAgeAccel.
- The rs17321515 A allele, associated with increased triglycerides^15^, is associated with increased BioAgeAccel, but decreased PhenoAgeAccel.
- The rs7412 T allele, or *APOE* e2 determined allele, associated with increased longevity^16^, is associated with decreased BioAgeAccel, but increased PhenoAgeAccel.

### Genetic Associations

Among the biomarkers, PhenoAge acceleration was genetically highly correlated with RDW (*r*_g_=0.65), followed by CRP (*r*_g_=0.48), and then white blood cell count (*r*_g_ = 0.46) (Figure S1). BioAge acceleration was genetically highly correlated with systolic blood pressure (*r*_g_ = 0.84), followed by alkaline phosphatase (*r*_g_=0.43), and then CRP (*r*_g_=0.36) (Figure S1).

The genetic correlation between PhenoAgeAccel and BioAgeAccel was 0.42 (SE=0.047). Both PhenoAgeAccel and BioAgeAccel had low genetic correlations with gastrointestinal diseases (GWAS summary statistics from Liu et al., 2015^17^), prostate and breast cancers^18,19^, and Alzheimer’s disease (Figure S2). Both were genetically correlated with coronary artery disease (CAD)^20^ (*r*_g_=0.27 with PhenoAgeAccel, *r*_g_=0.38 with BioAgeAccel), osteoarthritis^21^ (*r*_g_=0.30 with PhenoAgeAccel, *r*_g_=0.25 with BioAgeAccel), stroke^22^ (*r*_g_=0.30 with PhenoAgeAccel, *r*_g_=0.34 with BioAgeAccel), chronic kidney disease^23^ (*r*_g_=0.35 with PhenoAgeAccel, *r*_g_=0.26 with BioAgeAccel), type II diabetes^24^ (*r*_g_=0.36 with PhenoAgeAccel, *r*_g_=0.33 with BioAgeAccel), a 49-item frailty including pains and diseases^25^ (*r*_g_=0.34 with PhenoAgeAccel, *r*_g_=0.27 with BioAgeAccel), and parental mortality^26^ (*r*_g_=0.42 with PhenoAgeAccel, *r*_g_=0.45 with BioAgeAccel) (Figure S2).

PhenoAgeAccel (*r*_g_=0.44) was genetically more positively correlated with body mass index (BMI)^27^ than BioAgeAccel (*r*_g_=0.24). Waist circumstance and waist-hip ratio, adjusted for BMI and physical activity^28^, were not correlated with PhenoAgeAccel genetically, but a modest genetic correlation was found between BioAgeAccel and waist-hip ratio (*r*_g_=0.16). BioAgeAccel (systolic: *r*_g_ = 0.84, diastolic: *r*_g_=0.57) also was genetically more positively correlated with systolic and diastolic blood pressures than PhenoAgeAccel (systolic: *r*_g_=0.23, diastolic: *r*_g_=0.17). Genetically increased PhenoAgeAccel and BioAgeAccel were correlated with lower forced vital capacity (FVC)^27^ and forced expiratory volume in one second (FEV1) to a moderate degree^27^ but not with the FEV1/FVC ratio^27^. The genetic correlations between PhenoAgeAccel or BioAgeAccel with heel bone mineral density^27^ and heart rate variability (the root mean square of the successive differences of inter beat intervals, RMSSD)^29^ were minimal (Figure S3).

PhenoAgeAccel was genetically more correlated than BioAgeAccel with hematology traits^27^ with no surprise as four hematological measures are included in PhenoAge versus none in BioAge (Figure S4). PhenoAgeAccel and BioAgeAccel were genetically associated with different cholesterol biomarkers: total cholesterol, LDL cholesterol, and apolipoprotein B with BioAgeAccel, and HDL cholesterol and apolipoprotein A-1 with PhenoAgeAccel. Similarly, BioAgeAccel was genetically more correlated than PhenoAgeAccel with the liver biomarkers of alanine aminotransferase, aspartate aminotransferase, and gamma glutamyltransferase, whereas albumin, another liver biomarker, was more correlated with PhenoAgeAccel genetically. PhenoAgeAccel also was genetically more correlated than BioAgeAccel with creatinine, cystatin C, HbA1c, and CRP — biomarkers linked to kidney function, diabetes, and inflammation (Figure S5).

### Polygenic Risk Scores

5,198 SNPs (p<0.0064) were selected to calculate PRSs for PhenoAgeAccel, which explained 0.50% of the variance in PhenoAge, in addition to 74.23% by other covariates, primarily baseline chronological age, plus sex, baseline assessment center, genotyping array type, and the first five genetic principal components. Similarly, 146,223 SNPs (p<0.45) were selected for BioAgeAccel, accounting for 0.068% of the variance in BioAge, independent of 94.49% by baseline chronological age and other covariates. SNPs were selected for PRS to best explain the variance of PhenoAge or BioAge given other covariates were in the model. More SNPs were included in the PRS of BioAgeAccel than in the PRS of PhenoAgeAccel likely due to the small residual variance after accounting for other covariates and also small SNP effects in general for BioAgeAceel. While the variance independently explained by the PRS was minimal, the top 20% (high-risk class) and bottom 20% (low-risk class) of PRS showed distinct aging phenotypes.

The top 20% was compared to the bottom 20% of PhenoAgeAccel or BioAgeAccel PRS for a variety of aging traits adjusting for baseline chronological age, sex, baseline assessment center, genotyping array type, and the first five genetic principal components. The mean difference in PhenoAge between the top and bottom 20% of PhenoAgeAccel PRS (0.20 SD, 95% CI: 0.19 to 0.21 SD) was larger than the mean difference in BioAge between the top and bottom 20% of BioAge PRS (0.073 SD, 95% CI: 0.070 to 0.076 SD) in terms of either SD, PhenoAge SD=9.56 and BioAge SD=8.17. PhenoAge and BioAge share CRP, creatinine, and alkaline phosphatase in composition. Higher levels of the three biomarkers, particularly CRP, were observed in the top 20% than in the bottom 20% of PhenoAgeAccel or BioAgeAccel PRS (top left, Figure 4). The top-and-bottom mean difference of PhenoAgeAccel PRS was larger than that of BioAgeAccel PRS in biomarkers that appear in PhenoAge but not in BioAge, and vice versa (top left, Figure 4). Interestingly, the top 20% of PhenoAgeAccel PRS had lower mean cholesterol (−0.09 SD, 95% CI: −0.10 to −0.07 SD) than the bottom 20%, which was opposite for BioAgeAccel that the top 20% had 0.09 SD (95% CI: 0.08 to 0.11 SD) higher mean cholesterol than the bottom 20% (top left, Figure 4). The opposite trend was also found in mean corpuscular volume, with smaller top-and-bottom mean differences (top left, Figure 5).

**Figure 4.**
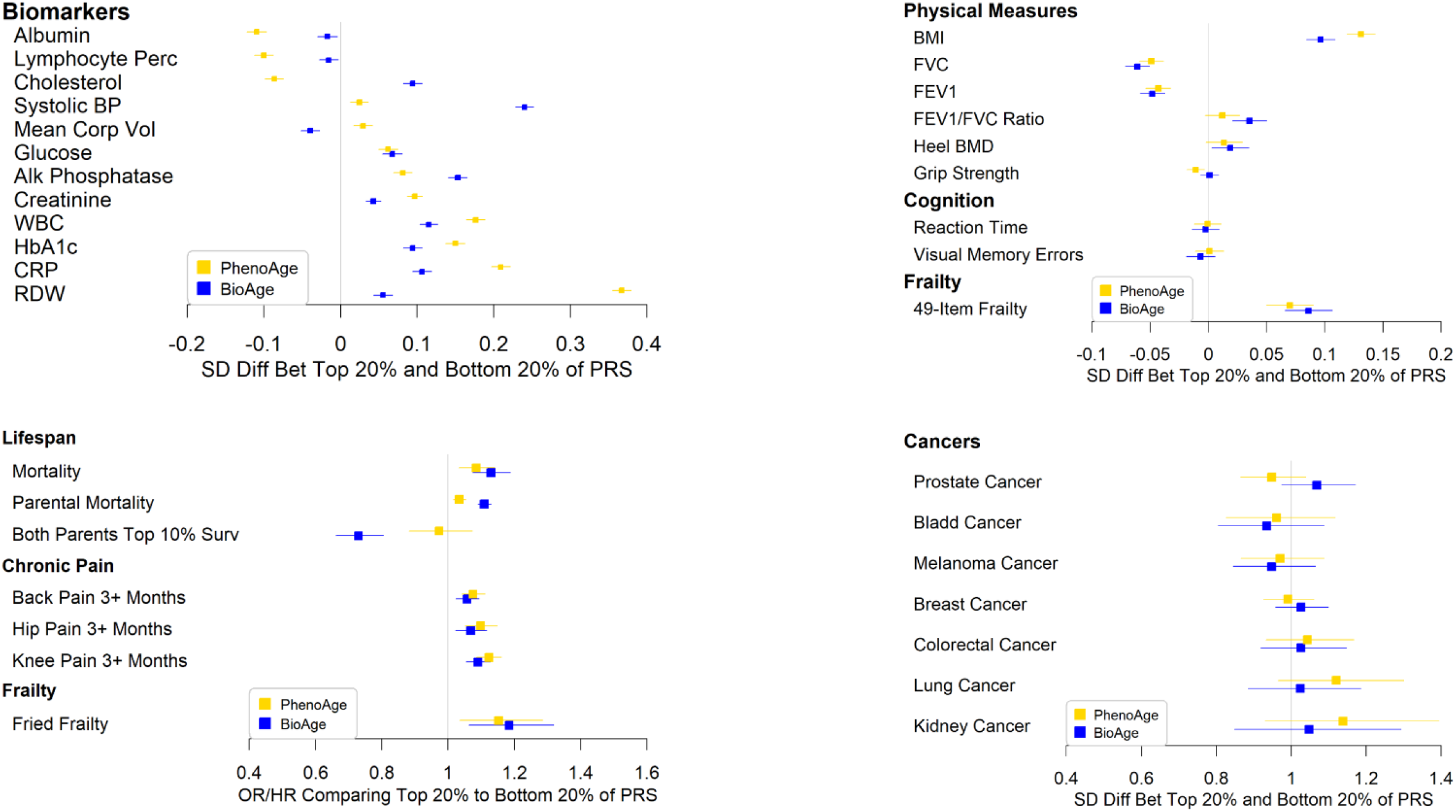
**Comparisons between the top 20% and bottom 20% of PhenoAgeAccel or BioAgeAccel PRS for biomarkers included in PhenoAge or BioAge and a variety of aging phenotypes**

**Figure 5.**
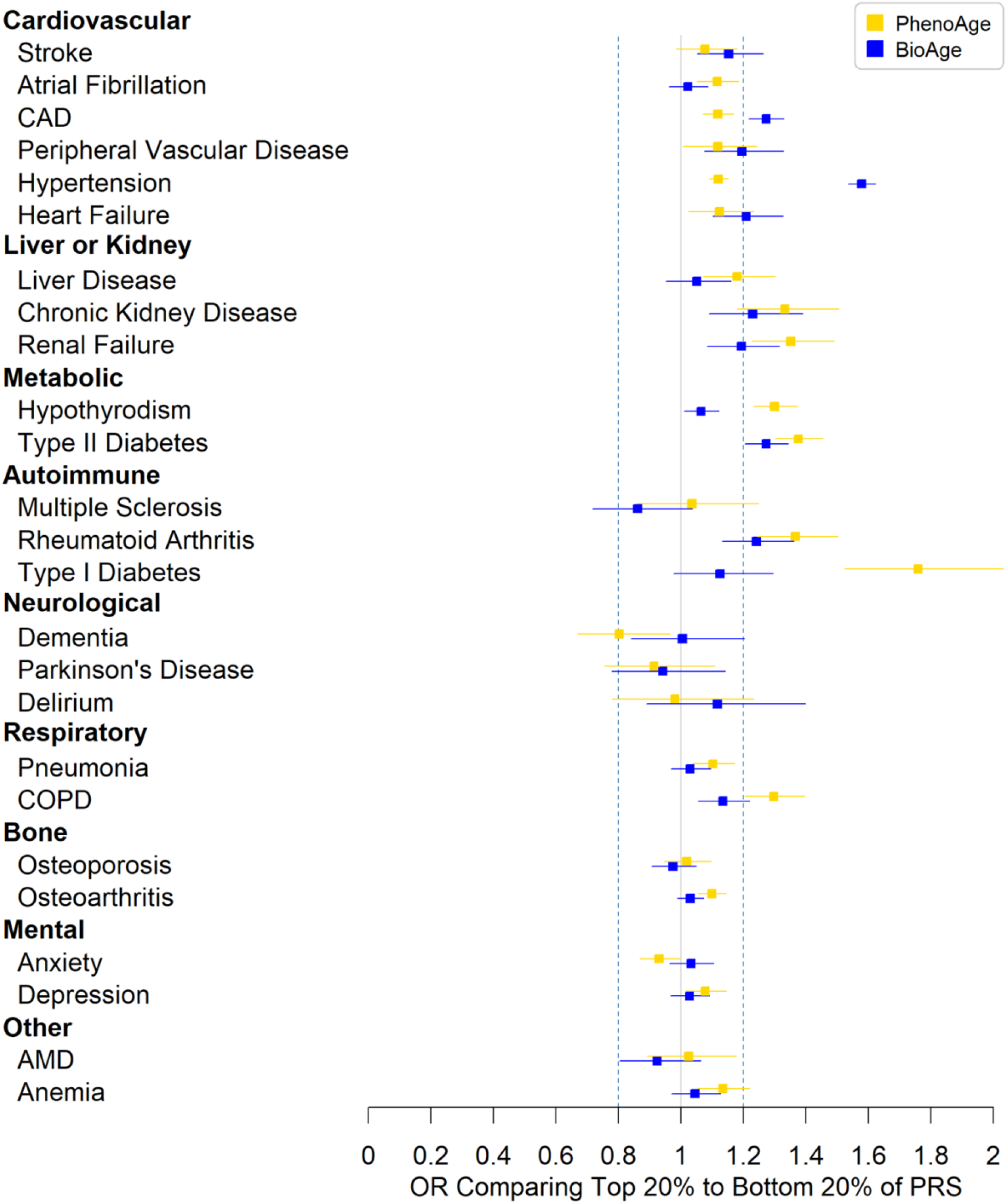
**Odds ratios (ORs) for diseases comparing the top 20% to the bottom 20% of PhenoAgeAccel or BioAgeAccel polygenic risk score (PRS)**

The top 20% of BioAgeAccel PRS were more likely to die early (HR=1.13, 95% CI: 1.07 to 1.19) and have higher parental mortality risk (HR = 1.11, 95% CI: 1.09 to 1.13), and were less likely to have both parents survive to the top 10% of sex-specific lifespans (OR = 0.73, 95% CI: 0.66 to 0.81) than the bottom 20%. Similar results were observed for PhenoAgeAccel PRS, but with smaller risk ratios, participant mortality HR=1.08 (95% CI: 1.03 to 1.14), parental mortality HR=1.03 (95% CI: 1.01 to 1.05), and parental longevity OR=0.97 (95% CI: 0.88 to 1.07) (bottom left, Figure 4). We also found higher likelihoods of chronic pain and Fried frailty^30^ (frail, if 3 or more items checked) for the top 20% versus the bottom 20% when considering either PhenoAgeAccel or BioAgeAccel PRS. The top 20% of PhenoAgeAccel or BioAgeAccel PRS was associated with higher mean BMI and more deficits in a 49-item frailty^31^ (a modified Rockwood frailty index, essentially accumulation of deficits^32^), plus lower FVC and FEV1 but not FEV1/FVC ratio, grip strength, heel bone mineral density, or cognitive measures of reaction time and visual memory errors (top right, Figure 4).

Both PhenoAgeAccel and BioAgeAccel PRSs were not associated with prevalent cancers including prostate cancer, breast cancer, and colorectal cancer (bottom right, Figure 4). The associations of BioAgeAccel PRS were stronger than those of PhenoAgeAccel PRS with prevalent cardiovascular diseases, particularly CAD and hypertension (Figure 5). The odds ratio of CAD was 1.27 (95% CI: 1.22 to 1.33) and that of hypertension was 1.58 (95% CI: 1.53 to 1.62) comparing the top 20% to the bottom 20% of BioAge PRS. At the biomarker levels, total cholesterol, low LDL cholesterol, apolipoprotein B, and triglycerides, risk factors of CAD, were elevated in the top 20% of BioAgeAccel PRS but reduced in the top 20% of PhenoAgeAccel PRS compared to the bottom 20% of each (Figure S6). These biomarker results suggested the association between PhenoAgeAccel PRS and CAD was likely driven by non-lipid mechanisms as indicated by the gene set analysis results.

PhenoAgeAccel PRS was more strongly associated than BioAgeAccel PRS with liver and kidney diseases (Figure 5) and the associated biomarkers, e.g. albumin, total bilirubin, creatinine, and cystatin C (Figure S6), plus COPD, hypothyroidism, type I and type II diabetes, and rheumatoid arthritis (Figure 5). The odds ratio of type I diabetes was 1.76 (95% CI: 1.52 to 2.03) and that of type II diabetes was 1.38 (95% CI: 1.30 to 1.45) comparing the top 20% to the bottom 20% of PhenoAgeAccel PRS, and those comparing the top 20% to the bottom 20% of BioAgeAccel PRS were 1.12 (95% CI: 0.98 to 1.30) for type I diabetes and 1.27 (95% CI: 1.20 to 1.34) for type II diabetes. The associations of PhenoAgeAccel or BioAgeAccel PRS were minimal with bone diseases such as osteoporosis and osteoarthritis, age-related macular degeneration (AMD), anxiety and depression, and two neurological disorders, Parkinson’s disease and delirium (Figure 5). A negative association was observed between PhenoAgeAccel PRS and dementia (OR=0.80, 95% CI: 0.67 to 0.96). This was mainly driven by *APOE*, which when adjusted for completely accounted for the association (OR=0.97, 95% CI: 0.80 to 1.16).

### Biological Age Measures and *APOE* Genotypes

Some of the strongest associations for both PhenoAgeAccel and BioAgeAccel were with *APOE* isoform coding SNPs, but the effect directions were opposite. The *APOE* e2 determined T allele of rs7412 was associated with increased PhenoAgeAccel but decreased BioAgeAccel. Similarly, the rs429358 C allele (*APOE* e4), a risk factor for Alzheimer’s disease, was associated with decreased PhenoAgeAccel but increased BioAgeAccel although the association with BioAgeAccel didn’t reach genome-wide significance (p=1.3×10^−7^).

Taking a step further, we associated PhenoAge and BioAge with *APOE* isoforms determined based on the genotypes of rs429358 and rs7412. For BioAge, results suggested that e2e3 and e2e2 were both associated with younger BioAge relative to the reference genotype (e3e3), while e3e4 and e4e4 exhibited higher BioAges, where the results were adjusted for baseline chronological age, sex, genotyping array type, baseline assessment center, and the first five genetic principal components (Figure 6). When comparing *APOE* genotypes as a function of PhenoAge, we find the reverse—e2e3 and e2e2 appeared older than e3e3, whereas e3e4, and e4e4 appeared younger.

**Figure 6.**
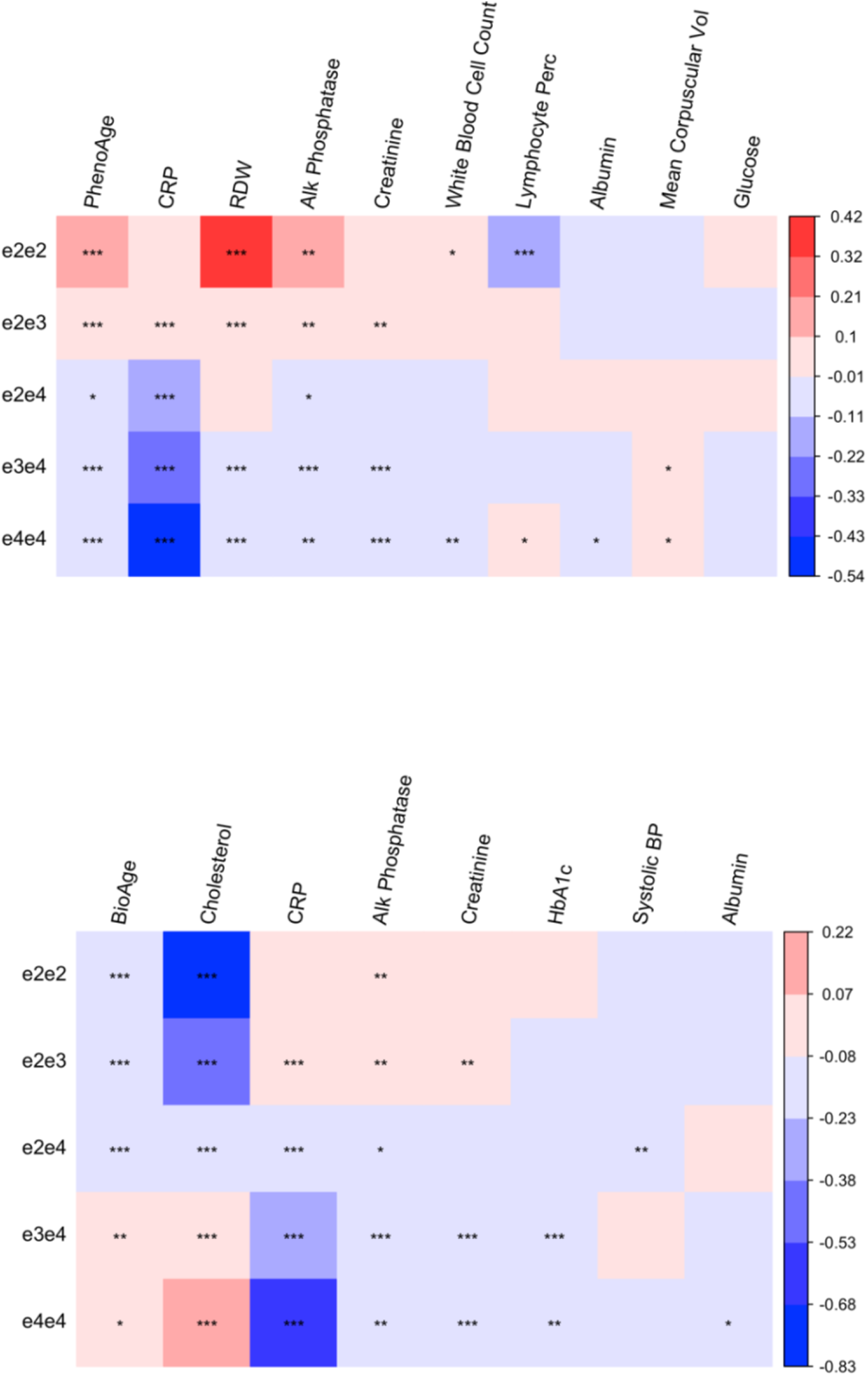
**Mean standard deviation (SD) differences between non-e3e3 and e3e3 genotypes: 1) biomarkers of PhenoAge (top) or BioAge (bottom) sorted by p-value from left to right for the null hypothesis of no genotypic effects; 2) p<0**.**05, p<0**.**01, and p<0**.**001 labelled by *, **, ***, respectively**.

To further disentangle the associations between *APOE* genotypes and accelerated aging by the two biological age measures, we examined the associations between *APOE* genotypes and the individual biomarkers that make up the composites. We found that the trend of mean BioAge (e4e4 > e4e3 > e3e3 > e2e4 > e2e3 > e2e2) also held for total cholesterol, which was most strongly associated with *APOE* genotypes among the biomarkers of BioAge (Figure 6). When adjusting for total cholesterol, the trend of mean BioAge was reversed, i.e., e3e4 and e4e4 younger than e2e3 and e2e2, suggesting that decelerated BioAge associated with e2 was driven by differences in plasma total cholesterol levels. The biomarkers that appeared to show inverse associations (similar to PhenoAge) were RDW, CRP, alkaline phosphatase, creatinine, and white blood cell count. For all of these biomarkers there was a trend towards higher levels among participants with e2 alleles and lower levels among those with e4 alleles (Figure 6).

## Discussion

Overall, our analysis using the UK Biobank biomarker data identified both overlapping and distinct genetic underpinnings of two widely applied biological age measures. Our results suggested that although the estimated heritability is similar for PhenoAgeAccel (14.45%) and BioAgeAccel (12.39%) with the genetic correlation being 0.42, these two measures capture distinct aging domains with different genetic determinants, as a result of their differential biomarker compositions. SNPs associated with BioAgeAccel (p<5×10^−8^) tended to relate to systolic blood pressure and lipid biomarkers, with enrichment analysis pointing to an increased proportion of genes involved in lipid homeostasis, plasma lipoprotein particle clearance, chylomicron, sterol homeostasis, and cholesterol transport activity. Conversely, SNPs associated with PhenoAgeAccel were shown to relate to CRP, white blood cell count, and RDW, and were enriched in biological processes involved in regulation of cell signaling by *CBL*, transcription, immune system process, and myeloid cell homeostasis.

The immune/inflammation versus lipid findings for PhenoAgeAccel and BioAgeAccel, respectively, were also recapitulated when comparing the associations between PRS and age-related outcomes. Results suggested that the top 20% of PhenoAgeAccel and BioAgeAccel PRS were differentially linked to a variety of diseases. For instance, BioAgeAccel PRS outperformed PhenoAgeAccel PRS in prioritizing cardiovascular and all-cause mortality risk in this young cohort, while PhenoAgeAccel PRS showed more robust associations than BioAgeAccel PRS for liver/kidney diseases, and chronic inflammatory and autoimmune diseases. The stronger link between BioAgeAccel PRS and all-cause mortality (compared to PhenoAgeAccel PRS) may be driven in part by its association with cardiovascular disease, which is the leading cause of death in the UK. By comparison, the diseases associated with PhenoAgeAccel PRS tend to contribute to major morbidity, while being less common causes of death. This may suggest that individuals genetically predisposed to accelerated BioAge may be more likely to experience shortened lifespan, while those genetically predisposed to accelerated PhenoAge, may not experience major reductions in lifespan, but may experience decreased healthspan (disease-free life expectancy). The hypothesis needs to be tested in older adults, however. Of note, accelerated aging is not only determined by genetics but also by environment. Interestingly, when considering the actual values rather than the PRS, accelerated PhenoAge is more strongly associated with all-cause mortality than accelerated BioAge in UK Biobank, which implies that the association between accelerated PhenoAge and all-cause mortality may be explained to a larger degree by the environmental components.

The PhenoAgeAccel PRS was also related to dementia, but in the opposite than the expected direction, such that individuals with increased PhenoAgeAccel had reduced odds of dementia. This result was almost entirely driven by the association between PhenoAgeAccel and *APOE*, which is the most well-known genetic risk factor for late-onset Alzheimer’s disease (LOAD). Our results suggested that while PhenoAgeAccel and BioAgeAccel were both associated with the two *APOE* isoform coding SNPs (rs429358 and rs7412), the relationships were inverse. For instance, the *APOE* e4 allele is traditionally associated with adverse health outcomes, including an increased risk of Alzheimer’s disease, cardiovascular disease, and reduced life expectancy, while the e2 allele confers protection. However, in our results, we observed increased PhenoAgeAccel associated with e2 genotypes and decreased PhenoAgeAccel associated with e4 genotypes, relative to the common e3e3 genotype. This paradoxical result was also found for a number of the biomarkers that make up PhenoAge, which likely explains this finding. For instance, *APOE* e2 allele was associated with higher CRP, RDW, alkaline phosphatase, creatinine, and white blood cell count, while *APOE* e4 allele was generally associated with lower levels of these biomarkers^33^. *APOE* e4 allele has previously been linked to lower CRP^34^; however, it remains unclear what drives the *APOE* e4-and-CRP association and the resulting consequence. Contrary to PhenoAgeAccel, BioAgeAccel showed an expected association with *APOE* that consisted of decelerated aging among participants with e2 alleles and accelerated aging among participants with e4 alleles. This association was accounted for by higher levels of total cholesterol among those with increased BioAgeAccel. This is in-line with *APOE* known function as a transporter of extracellular cholesterol and the existing evidence suggesting those with the e2 allele exhibit reduced circulating cholesterol, particularly low-density lipoproteins (LDL)^33^.

Inevitably, our study has limitations. The UK Biobank participants are healthier than the general population^35^; therefore, are less susceptible to accelerated aging. The disease status was determined based on self-reported doctor diagnoses at baseline and electronic health records to 2017. Given that some participants were still relatively young and will likely go on to develop late-onset morbidity this will contribute to misclassification, which could bias associations towards the null. Nevertheless, when disease prognostic biomarkers were analyzed, we observed consistent results. Last but not least, our findings are based on European-descent participants and may not be generalizable to other ancestry populations.

Overall, the mapped genes and enriched genes sets highlight that these two biological age measures may capture different aspects of the aging process—cardiometabolic by BioAge and inflammaging/immunoscenece by PhenoAge. Nevertheless, PhenoAgeAccel and BioAgeAccel PRSs are not disease-specific and can be used to prioritize genetic risk for multiple morbidity or mortality outcomes—particularly cardiovascular diseases and all-cause mortality via BioAge, and liver or kidney diseases, COPD, rheumatoid arthritis, hypothyroidism, and type I and type II diabetes via PhenoAge. Our findings confirm the hypothesis that individuals may age in different ways, due in part to different underlying genetic susceptibility. In moving forward, understanding personalized aging susceptibility phenotypes has important implications for primary and secondary disease interventions.

## Methods

### UK Biobank

Over 500,000 participants between the ages of 40 and 70 were recruited by UK Biobank from 2006 to 2010 ^36,37^, of which, over 90% of the cohort were European-descent. Phenotypes considered in this study include participant mortality, parental lifespan, cognitive function, physical measures, and diseases. The death status was determined based on death certificate data, updated to March 2020 for all participants. Some deaths were recorded in April 2020 but the mortality data for that month is incomplete. The disease diagnosis was confirmed based on self-reported doctor diagnoses at baseline, cancer registry data to 2016, and hospital admission records from 1996 to 2017. A list of disease ICD-10 codes used to identify diseases is provided in Table S4. At recruitment, participants completed online questionnaire and physical measurements and their biological samples were collected for biomarker assays. Physical measurements were described elsewhere^33^. A full list and technical details are available via the UK Biobank Biomarker Panel^38^ and UK Biobank Haematology Data Companion Document^39^.

### Genetic Data

DNA was extracted from blood samples and was genotyped using Affymetrix UK BiLEVE Axiom array for the first ~50,000 participants and Affymetrix UK Biobank Axiom array for the remaining cohort – the two arrays have over 95% overlap ^37^. Imputation was performed by the UK Biobank team using the reference panels of 1000 Genomes and the Haplotype Reference Consortium (HRC), yielding ~93 million variants in 487,442 participants. Of whom, participant (n=968) with unusually high heterozygosity or missing genotype calls were further removed^37^.

### Biological Age Measures

Biomarkers included in PhenoAge^1^ and/or BioAge^2^ are listed in Table 1. To correct distribution skewness, we set the bottom 1% of values to the 1^st^ percentile and the top 1% to the 99^th^ percentile. PhenoAge was developed based on mortality scores from the Gompertz proportional hazard model on chronological age and nine biomarkers, which were selected from 42 clinical biomarkers by Cox penalized regression model predicting age-related mortality in the NHANES III^1^. The formula of PhenoAge is given by

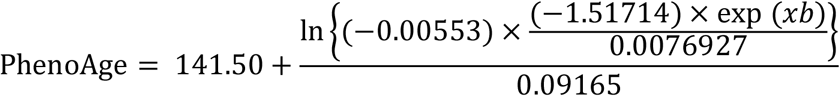

where

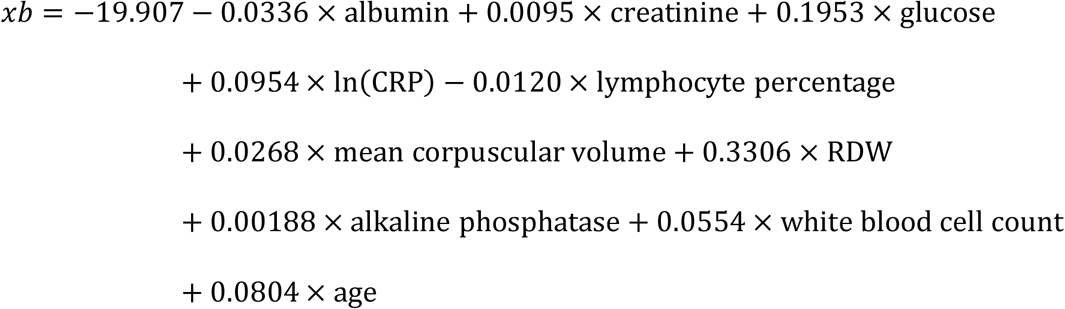

and *age* denotes the chronological age.

Seven biomarkers and chronological age were used to calculate BioAge, where albumin, creatinine, CRP, and alkaline phosphatase were overlapped between PhenoAge and BioAge. BioAge^2^ was trained for the biological age surrogate of chronological age, using the NHANES III data by applying an algorithm previously proposed by Klenmera and Doubal^40^,

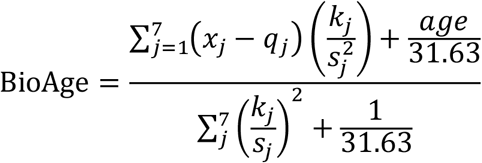

where *x*_*j*_ denotes the level of *j*-th biomarker, with the corresponding parameters *q*_*j*_, *k*_*j*_, and *s*_*j*_ provided in Table S5. *age* here, again, denotes the chronological age. Biological age acceleration was estimated by the residual of PhenoAge or BioAge after subtracting the effect of chronological age using a linear regression model, termed PhenoAgeAccel and BioAgeAccel, respectively.

### Included Samples

Participants of European descent were included, identified using genetic principal components analysis in detail in Thompson and colleagues ^41^. Additionally, one in third-degree or closer pairs were removed, identified via pairwise kinship coefficients. The sample was randomly split into a training and a testing set, with a 1 to 2 ratio. The training set was used to perform genome-wide association analysis with the results being used to create PRSs in the testing set to evaluate the use for risk stratification for age-related outcomes.

### SNP Quality Control

Of 93,095,623 genotyped or imputed SNPs, 16,446,666 SNPs passed the quality control, where SNPs were excluded if meeting any of the criteria: 1) imputation information score <0.3, 2) minor allele frequency <0.1%, 3) Hardy-Weinberg equilibrium test p-value significant at the Bonferroni-corrected level, 4) missing imputation information score, minor allele frequency, or Hardy-Weinberg equilibrium test result. The SNP summary statistics were calculated using the QCTOOL software version 2^42^.

### Genome-Wide Association Analysis

The association between accelerated PhenoAge or BioAge with each SNP was examined using an efficient Bayesian linear mixed effects model (BOLT-LMM software version 2.2)^43^ for the outcome of PhenoAge or BioAge with additive allelic effect of the candidate SNP, and other fixed effects: chronological age (to make the case of accelerated biological age), sex, genotyping array type, and assessment center, plus random polygenic and environment effects. By default, the LD scores included in the BOLT-LMM for European-ancestry samples were used to calibrate the BOLT-LMM statistic. SNP p-values smaller than 5×10^−8^ were deemed to be statistically significant. Manhattan plots were created for visualization using the FUMA (Functional Mapping and Annotation) software version 1.3.5^44^. The genomic inflation due to population stratification or cryptic relatedness was evaluated by linkage disequilibrium (LD) score regression^45^, where SNPs were filtered to the HapMap3 SNPs, well imputed in most studies to avoid bias from poor imputation quality. The LD scores were downloaded from the url (https://data.broadinstitute.org/alkesgroup/LDSCORE/), precomputed using the European data from the 1000 genome project phase 3.

We performed a stepwise model selection procedure on the genome-wide SNP summary statistics to identify independent signals (p<5×10^−8^) using the COJO (Conditional & JOint association analysis) function in the GCTA (Genome-wide Complex Trait Analysis) software^46^ version 1.92.1 beta6 Linux. SNPs more than 10,000 kb away from each other were assumed to be in complete linkage equilibrium. As SNPs were selected, those with multiple regression R^2^ greater than 0.9 with already pre-selected SNPs were excluded, so as not to include redundant signals from high LD. The loci marked by the selected SNPs were mapped to genes based on GRCh37/hg19 coordinates, and were used in searches for published GWAS associations based on GWAS catalog^11^.

### Gene Enrichment Analysis

The GWAS p-values were analyzed by Multi-marker Analysis of GenoMic Annotation (MAGMA)^47^ in FUMA to perform a comparative gene-set analysis to test if genes in the gene set were more strongly associated with PhenoAgeAccel or BioAgeAccel than others, for 10,678 gene sets (curated gene sets: 4,761, GO terms: 5,917) from the MsigDB v6.2^48,49^. Additionally, a gene-property analysis was performed to test for positive relationships (one-sided test) between tissue-specific gene expression profiles and gene associations with PhenoAgeAccel or BioAgeAccel, using 53 tissue types from the GTEx repository version^50^. Both test results were adjusted for multiple testing using the Bonferroni correction method.

### Genetic Correlations

Genetic correlations of PhneoAgeAccel or BioAgeAccel were calculated by LD score regression^45^ using GWAS summary statistics, filtered to HapMap3 SNPs. GWAS summary statistics were downloaded from previous published GWAS. Those of biomarkers, not limited to PhenoAge or BioAge biomarkers, were downloaded from the Neale Lab^27^, where biomarkers were transformed by the rank-based inverse normal transformation, and the SNP-biomarker associations were adjusted for age, age^2^, sex, age × sex, age^2^ × sex and the top 20 genetic principal components in over 361,000 UK Biobank participants.

### Polygenic Risk Scores

The PRSice-2 software version 2.2.2^51^ was used to perform polygenic risk score (PRS) analysis. SNPs were clumped to obtain SNPs in low LD (*r*_2_<0.1) in a 250 base-pair window. SNPs with p-values smaller than a threshold were used to calculate the PRS, sum of the effect alleles associated with accelerated aging, weighted by the effect size. The optimal threshold was chosen by a grid search from 1×10^−5^ to 0.5 with the increment of 1×10^−5^ plus 1, such that the variance of PhenoAge or BioAge in the testing set was best explained by PRS, in addition to that by baseline chronological age, sex, genotyping array type, baseline assessment center, and the first five genetic principal components. Subjects were equally divided into five groups by the PRS, where the top 20% (high-risk class) was compared to the bottom 20% (low-risk class) for a variety of aging traits. The association analysis was conducted using a regression model, with adjustment for baseline chronological age, sex, genotyping array type, baseline assessment center, and the first five genetic principal components.

## Data Availability

This research was conducted using the UK Biobank resource, under the application 14631.

https://doi.org/10.6084/m9.figshare.12620291.v1

https://doi.org/10.6084/m9.figshare.12620366.v1

## Data Availability

The GWAS summary statistics for PhenoAgeAccel and BioAgeAccel can be downloaded at figshare:

PhenoAgeAccel GWAS summary statistics https://doi.org/10.6084/m9.figshare.12620291.v1

BioAgeAccel GWAS summary statistics https://doi.org/10.6084/m9.figshare.12620366.v1

## Acknowledgements

CLK and MEL are supported in part by a R00 grant (R00AG052604) funded by the National Institute on Aging, National Institute Health, USA. CLK and LCP are supported in part by a R21 grant (R21AG060018) funded by the National Institute on Aging, National Institute Health, USA. LCP is supported by the University of Exeter Medical School, and in part by the University of Connecticut School of Medicine. JLA is supported by UK Medical Research Council award (MR/S009892/1). UK Biobank received an approval from the UK Biobank Research Ethics Committee (REC; REC reference 11/NW/0382). All the participants provided written informed consent to participate in the study and for their data to be used in future research. This research was conducted using the UK Biobank resource, under the application 14631. The views expressed in this publication are those of the author(s) and not necessarily those of the NHS, the National Institute for Health Research or the Department of Health and social care.

## Author Contributions

Study design: CLK, MEL; data analysis: CLK; manuscript preparation: all

## Competing Interests

None

## Notes

### Competing Interest Statement

The authors have declared no competing interest.

### Author Declarations

Project is not human subject research and IRB involvement is not required.

